# Measurement of SARS-CoV-2 antigens in plasma of pediatric patients with acute COVID-19 or MIS-C using an ultrasensitive and quantitative immunoassay

**DOI:** 10.1101/2021.12.08.21267502

**Authors:** George B. Sigal, Tanya Novak, Anu Mathew, Janet Chou, Yubo Zhang, Navaratnam Manjula, Predeepthi Bathala, Jessica Joe, Nikhil Padmanabhan, Daniel Romero, Gabriella Allegri-Machado, Jill Joerger, Laura L. Loftis, Stephanie P. Schwartz, Tracie C. Walker, Julie C. Fitzgerald, Keiko M. Tarquinio, Matt S. Zinter, Jennifer E. Schuster, Natasha B. Halasa, Melissa L. Cullimore, Aline B. Maddux, Mary A. Staat, Katherine Irby, Heidi R. Flori, Bria M. Coates, Hillary Crandall, Shira J. Gertz, Adrienne G. Randolph, Nira R. Pollock, on behalf of the Overcoming COVID-19 Investigators

**Affiliations:** Meso Scale Diagnostics, LLC., Rockville, MD, USA; Department of Anesthesiology, Critical Care and Pain Medicine, Boston Children’s Hospital and Department of Anesthesia, Harvard Medical School, Boston, MA, USA; Department of Pediatrics, Boston Children’s Hospital and Harvard Medical School, Boston, MA, USA; Institutional Centers for Clinical and Translational Research, Boston Children’s Hospital, Boston, MA, USA; Department of Laboratory Medicine, Boston Children’s Hospital, Boston, MA USA; Division of Critical Care Medicine, Department of Pediatrics, Baylor College of Medicine, Houston, TX, USA; Department of Pediatrics, University of North Carolina at Chapel Hill Children’s Hospital, Chapel Hill, NC, USA; Division of Critical Care, Department of Anesthesiology and Critical Care, The University of Pennsylvania Perelman School of Medicine, Philadelphia, PA, USA; Division of Critical Care Medicine, Department of Pediatrics, Emory University School of Medicine, Children’s Healthcare of Atlanta, Atlanta, GA, USA; Department of Pediatrics, Divisions of Critical Care and Bone Marrow Transplantation, University of California, San Francisco, San Francisco, CA, USA; Division of Pediatric Infectious Diseases, Department of Pediatrics, Children’s Mercy Kansas City, Kansas City MO, USA; Division of Pediatric Infectious Diseases, Department of Pediatrics, Vanderbilt University Medical Center, Nashville, TN, USA; Department of Pediatrics, College of Medicine, University of Nebraska Medical Center, Omaha, NE, USA; Department of Pediatrics, Section of Critical Care Medicine, University of Colorado School of Medicine and Children’s Hospital Colorado, Aurora, CO, USA; Department of Pediatrics, University of Cincinnati, Division of Infectious Diseases, Cincinnati Children’s Hospital Medical Center, Cincinnati, OH, USA; Section of Pediatric Critical Care, Department of Pediatrics, Arkansas Children’s Hospital, Little Rock, AR, USA; Division of Pediatric Critical Care Medicine, Department of Pediatrics, Mott Children’s Hospital and University of Michigan, Ann Arbor, MI, USA; Division of Critical Care Medicine, Department of Pediatrics, Northwestern University Feinberg School of Medicine, Ann & Robert H. Lurie Children’s Hospital of Chicago, Chicago, IL, USA; Division of Pediatric Critical Care, Department of Pediatrics, University of Utah, Salt Lake City, UT, USA; Division of Pediatric Critical Care, Department of Pediatrics, Saint Barnabas Medical Center, Livingston, NJ, USA; Department of Laboratory Medicine, Boston Children’s Hospital and Harvard Medical School, Boston, MA, USA

**Keywords:** SARS-CoV-2, COVID-19, antigen, ultrasensitive immunoassay, antigenemia

## Abstract

**Background:** Detection of SARS-CoV-2 antigens in blood has high sensitivity in adults with acute COVID-19, but sensitivity in pediatric patients is unclear. Recent data suggest that persistent SARS-CoV-2 spike antigenemia may contribute to multisystem inflammatory syndrome in children (MIS-C). We quantified SARS-CoV-2 nucleocapsid (N) and spike (S) antigens in blood of pediatric patients with either acute COVID-19 or MIS-C using ultrasensitive immunoassays (Meso Scale Discovery).

**Methods:** Plasma was collected from inpatients (<21 years) enrolled across 15 hospitals in 15 US states. Acute COVID-19 patients (n=36) had a range of disease severity and positive nasopharyngeal SARS-CoV-2 RT-PCR within 24 hours of blood collection. Patients with MIS-C (n=53) met CDC criteria and tested positive for SARS-CoV-2 (RT-PCR or serology). Controls were patients pre-COVID-19 (n=67) or within 24h of negative RT-PCR (n=43).

**Results:** Specificities of N and S assays were 95-97% and 100%, respectively. In acute COVID-19 patients, N/S plasma assays had 89%/64% sensitivity, respectively; sensitivity in patients with concurrent nasopharyngeal swab cycle threshold (Ct) ≤ 35 were 93%/63%. Antigen concentrations ranged from 1.28-3,844 pg/mL (N) and 1.65-1,071 pg/mL (S) and correlated with disease severity. In MIS-C, antigens were detected in 3/53 (5.7%) samples (3 N-positive: 1.7, 1.9, 121.1 pg/mL; 1 S-positive: 2.3 pg/mL); the patient with highest N had positive nasopharyngeal RT-PCR (Ct 22.3) concurrent with blood draw.

**Conclusions:** Ultrasensitive blood SARS-CoV-2 antigen measurement has high diagnostic yield in children with acute COVID-19. Antigens were undetectable in most MIS-C patients, suggesting that persistent antigenemia is not a common contributor to MIS-C pathogenesis.

**Key points:** In a U.S. pediatric cohort tested with ultrasensitive immunoassays, SARS-CoV-2 nucleocapsid antigens were detectable in most patients with acute COVID-19, and spike antigens were commonly detectable. Both antigens were undetectable in almost all MIS-C patients.

## Introduction

Severe acute respiratory syndrome coronavirus 2 (SARS-CoV-2) infection in pediatric populations can present with a range of disease severity, from asymptomatic infection and mild illness to severe systemic disease with involvement of multiple organ systems [1]. Multisystem inflammatory syndrome in children (MIS-C) is a relatively rare sequela of SARS-CoV-2 infection initially recognized in early 2020, when otherwise healthy children were hospitalized with severe systemic inflammation, with timing consistent with a post-infectious syndrome [2].

Diagnosis of acute SARS-CoV-2 infection in children is similar to diagnosis in adults, including detection of viral RNA by nucleic acid amplification testing (NAAT) or viral antigens by immunoassay testing of upper respiratory specimens. Both types of measurements are likely to act as relative indicators of viral load, and nucleocapsid (N) antigen concentrations in nasopharyngeal (NP) swab samples have been shown to correlate closely with NAAT cycle threshold (Ct) values [3, 4]. In contrast, MIS-C is a syndrome defined by clinical criteria (including fever, inflammation, severity requiring hospitalization and multisystem involvement) plus evidence of current or recent SARS-CoV-2 infection (by NAAT, serology or antigen test; or exposure in past 4 weeks), with no alternative diagnosis [5]. Some patients with multisystem involvement meet clinical criteria for both severe acute COVID-19 and MIS-C [1].

Recent advances in SARS-CoV-2 diagnostic approaches include assays developed using two ultrasensitive and quantitative antigen detection technologies [Single Molecule Array (Simoa) (Quanterix, Billerica, MA) and S-PLEX® electrochemiluminescence immunoassay (MesoScale Discovery, Rockville, MD)] for use in both respiratory and non-respiratory specimens. Simoa and S-PLEX assays have detected N antigens with high sensitivity in the blood of adults with acute COVID-19 [6-8]. Data on the clinical performance of these assays for diagnosing children with either acute COVID-19 or MIS-C are limited. A recent study using Simoa-based assays concluded that SARS-CoV-2 spike (S) antigens were detectable in the blood of children with MIS-C. This finding raised the question of whether antigen detection could provide diagnostic utility in MIS-C and generated hypotheses about possible disease mechanism and therapeutic approaches based on a potential intestinal source of antigen leakage [9].

In this study, we applied ultrasensitive and quantitative S-PLEX assays for SARS-CoV-2 N [3, 4, 8, 10] and S antigens to plasma of hospitalized pediatric patients with either acute COVID-19 or MIS-C enrolled in a large multi-site study comparing the two presentations. We sought to characterize the range of SARS-CoV-2 antigen concentrations in blood of children with acute COVID-19 or MIS-C, further clarifying diagnostic options for two important presentations of pediatric COVID-19.

## Methods

### Clinical Cohorts and Sample Collection

COVID-19 Acute and COVID-19 MIS-C samples were collected in the Overcoming COVID-19 Immunobiology Study that investigates severe pediatric complications related to COVID-19 [2]. Samples were collected between June 17, 2020, and June 17, 2021 across 15 pediatric hospital sites in 15 U.S. states. Sites relied on the Boston Children’s Hospital IRB; informed consent was obtained from at least one parent or legal guardian. Patients were approached for enrollment and research sample collection as soon as possible after admission. Patients were classified as having acute COVID-19 if they had a positive SARS-CoV-2 reverse transcription polymerase chain reaction (RT-PCR) test and symptoms consistent with COVID-19 (Supplementary Methods). Only patients with acute COVID-19 that had a research NP swab sample collected within 24 hours of the research blood sample were included. The research NP sample was frozen and tested by RT-PCR at Vanderbilt University Medical Center (Halasa Lab) using the CDC EUA protocol (https://www.fda.gov/media/134922/download). N1 and N2 target Ct values were averaged for analysis. All MIS-C patients met CDC diagnostic criteria and were required to have a positive SARS-CoV-2 test (Supplementary Methods) [2, 5]. For all MIS-C patients, the RT-PCR result associated with each research blood sample was the most recent (preceding) clinical RT-PCR result reported by the hospital site. Clinical Ct values were not available. Some MIS-C patients also had a research NP swab collected within 24 hours of the research blood, which was tested by RT-PCR at Vanderbilt. Additional details are provided in Supplementary Methods.

Pre-COVID-19 control samples were discarded heparin plasma samples from pediatric patients (aged ≤18 years) with suspected *Clostridioides difficile* infection or discarded EDTA plasma samples from pediatric patients with suspected sepsis, captured prior to December 2019 under separate IRB protocols.

COVID-19 negative control samples were discarded heparin plasma samples from symptomatic and asymptomatic pediatric patients (aged ≤18 years) who had tested negative on SARS-CoV-2 RT-PCR testing of a respiratory sample collected on the same date (April 25-May 3, 2021). Samples were frozen within 24 hours of initial collection.

### SARS-CoV-2 Antigen and Serologic Assays

Detection of SARS-CoV-2 nucleocapsid (N) and spike (S) proteins was performed using MSD® S-PLEX CoV-2 N and MSD S-PLEX CoV-2 S assay kits (Meso Scale Discovery, Rockville, MD). The assays were run according to protocols in the kit package inserts [11, 12]. Plasma samples were diluted 4-fold in assay buffer prior to analysis. Sample quantitation was achieved using a calibration curve generated using a recombinant antigen standard. For graphing and analysis, any concentrations below the limit of detection (LOD) were assigned the LOD value, and any concentrations above the highest calibration standard were assigned its value. The LOD and assay cut-off for the N assay were 0.64 and 1.28 pg/mL, respectively, and for the S assay were 1.12 and 1.65 pg/mL, respectively (assay details in Supplementary Methods).

All samples were also tested using an MSD multiplexed serologic assay that measured IgG antibodies against SARS-CoV-2 N, S, and the spike receptor binding domain (RBD) and N-terminal domain (NTD), as well as antibodies against S from SARS-CoV-1 and common circulating coronaviruses (229E, HKU1, NL63 and OC43). Details of this MSD antibody panel are in Supplementary Methods. The assays were run according to the protocol provided with the assay kits [13].

Statistical methods are detailed in Supplementary Methods.

## Results

### Measurement of SARS-CoV-2 Antigen Levels in Pediatric Plasma

**Table 1** summarizes patient demographics and clinical data for acute COVID-19 patients (n=36; age range: 0.1-20.8 years; 22% previously healthy) and MIS-C patients (n=53; age range: 1.0-19.1 years; 79% previously healthy). **Table S1** summarizes key laboratory and blood sample handling data. **Figure 1** shows measured concentrations of N and S antigens in plasma for the four categories of study patients: pre-COVID-19 controls (“Pre-COVID-19,” n=67), RT-PCR-negative (ruled out) controls (“COVID-19 Negative,” n=43), acute COVID-19 cases (“COVID-19 Acute,” n=36) and MIS-C cases (“COVID-19 MIS-C,” n=53). As expected, antigen concentration measurements for the two negative control categories were low; only four (3.6%) samples (all N measurements) were slightly above the assay cut-offs. N and S antigen concentrations in acute COVID-19 cases (all with positive RT-PCR results on admission) spanned a wide range: <1.28 pg/mL (assay cut-off value) to >3,844 pg/mL (top of the calibration curve) for N, and <1.65 pg/mL (assay cut-off value) to 1,071 pg/mL for S. Two of the 36 acute COVID-19 patients had received intravenous immunoglobulin (IVIG) prior to blood collection; their N/S concentrations were 1024.8/8.65 pg/mL and 3844.0/1071.2 pg/mL, respectively, suggesting that IVIG did not inhibit antigen detection (the first patient also received monoclonal antibody treatment pre-sampling).

**Table 1:**
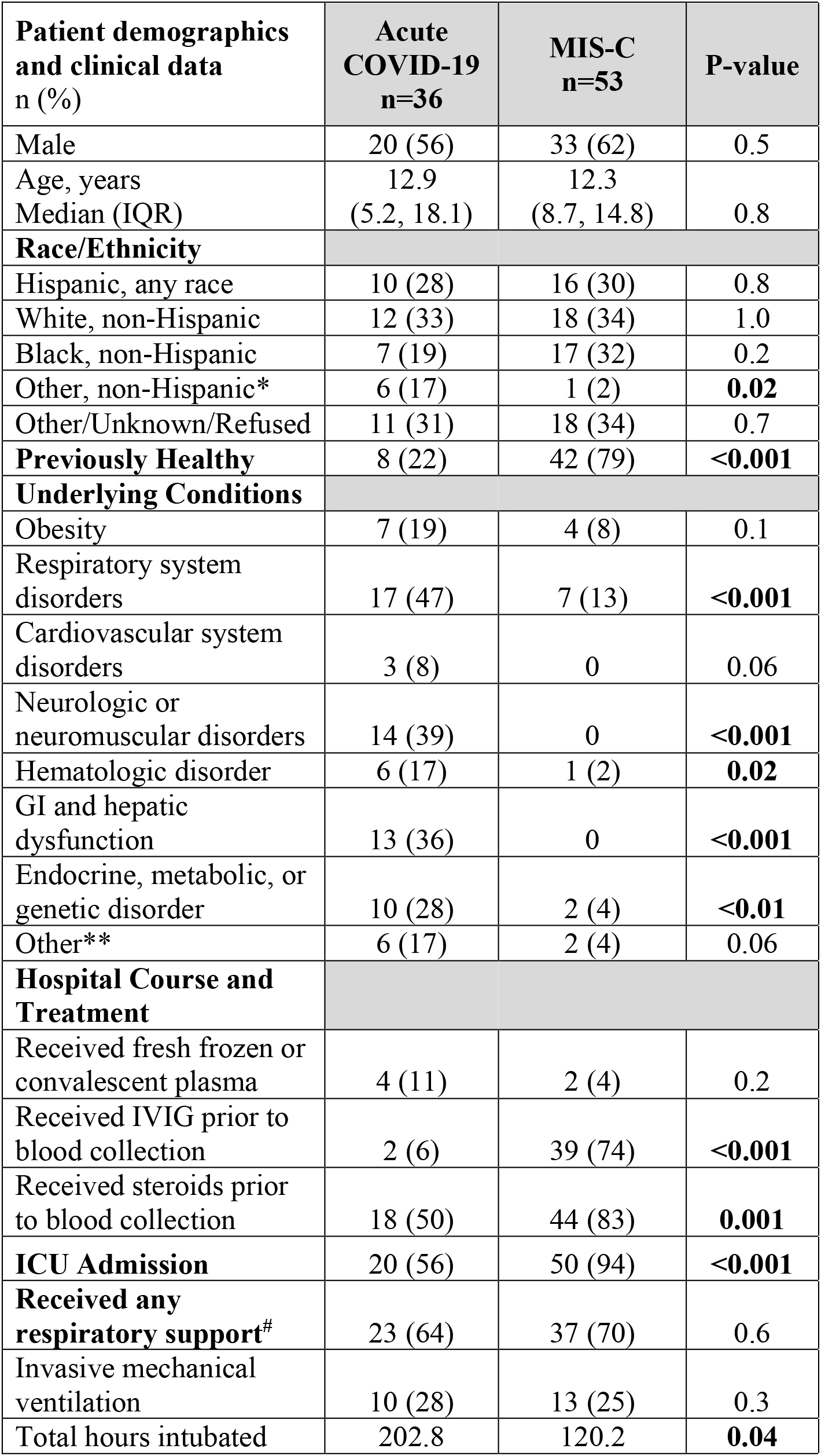
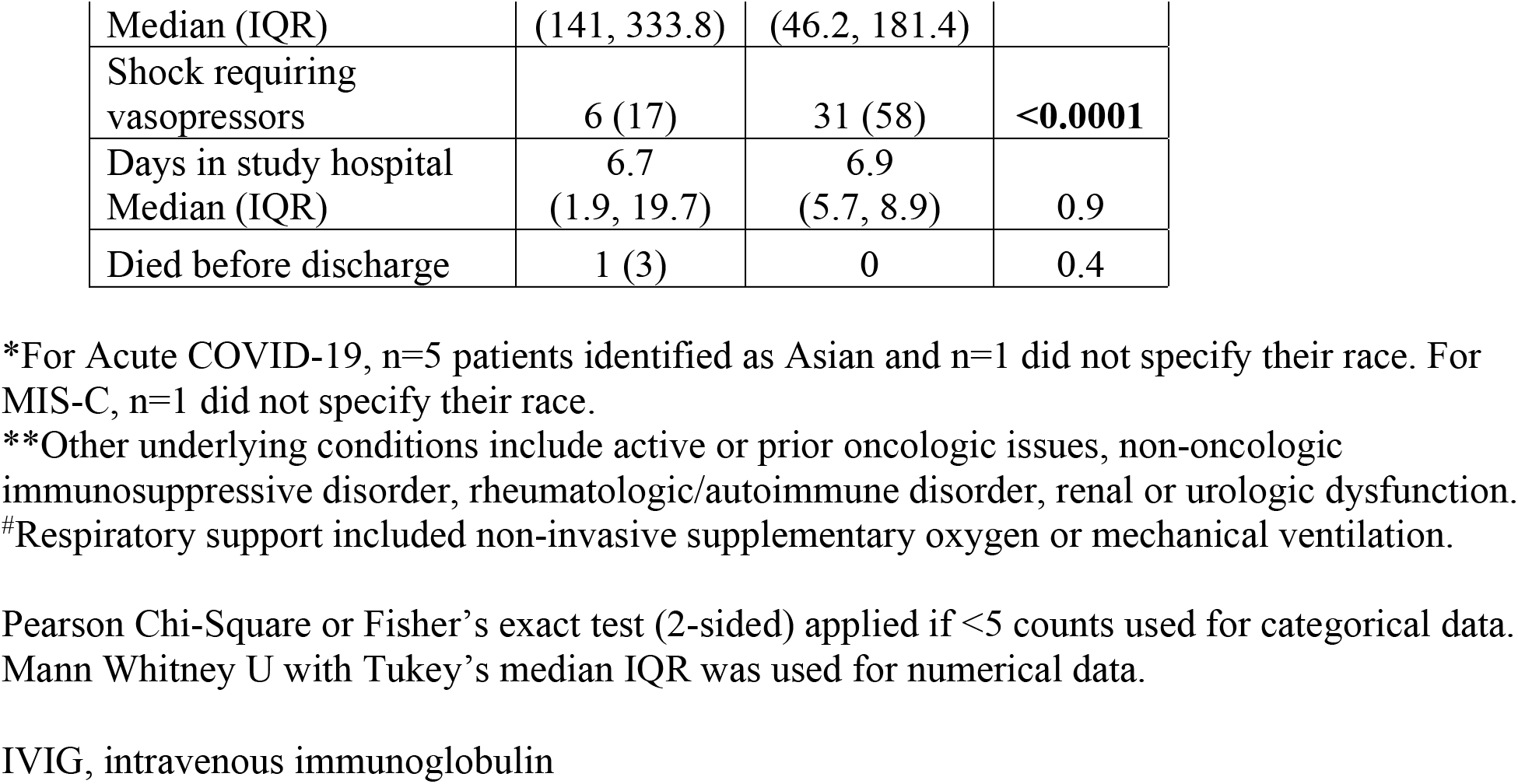
Patient demographics and clinical data.

| **Patient demographics and clinical data**<br>n (%) | **Acute COVID-19**<br>**n=36** | **MIS-C**<br>**n=53** | **P-value** |
| --- | --- | --- | --- |
| Male | 20 (56) | 33 (62) | 0.5 |
| Age, years | 12.9 | 12.3 |  |
| Median (IQR) | (5.2, 18.1) | (8.7, 14.8) | 0.8 |
| **Race/Ethnicity** |  |  |  |
| Hispanic, any race | 10 (28) | 16 (30) | 0.8 |
| White, non-Hispanic | 12 (33) | 18 (34) | 1.0 |
| Black, non-Hispanic | 7 (19) | 17 (32) | 0.2 |
| Other, non-Hispanic* | 6 (17) | 1 (2) | **0.02** |
| Other/Unknown/Refused | 11 (31) | 18 (34) | 0.7 |
| **Previously Healthy** | 8 (22) | 42 (79) | **<0.001** |
| **Underlying Conditions** |  |  |  |
| Obesity | 7 (19) | 4 (8) | 0.1 |
| Respiratory system disorders | 17 (47) | 7 (13) | **<0.001** |
| Cardiovascular system disorders | 3 (8) | 0 | 0.06 |
| Neurologic or neuromuscular disorders | 14 (39) | 0 | **<0.001** |
| Hematologic disorder | 6 (17) | 1 (2) | **0.02** |
| GI and hepatic dysfunction | 13 (36) | 0 | **<0.001** |
| Endocrine, metabolic, or genetic disorder | 10 (28) | 2 (4) | **<0.01** |
| Other** | 6 (17) | 2 (4) | 0.06 |
| **Hospital Course and Treatment** |  |  |  |
| Received fresh frozen or convalescent plasma | 4 (11) | 2 (4) | 0.2 |
| Received IVIG prior to blood collection | 2 (6) | 39 (74) | **<0.001** |
| Received steroids prior to blood collection | 18 (50) | 44 (83) | **0.001** |
| **ICU Admission** | 20 (56) | 50 (94) | **<0.001** |
| **Received any respiratory support<sup>#</sup>** | 23 (64) | 37 (70) | 0.6 |
| Invasive mechanical ventilation | 10 (28) | 13 (25) | 0.3 |
| Total hours intubated | 202.8 | 120.2 | **0.04** |
| Median (IQR) | (141, 333.8) | (46.2, 181.4) |  |
| Shock requiring vasopressors | 6 (17) | 31 (58) | **<0.0001** |
| Days in study hospital Median (IQR) | 6.7<br>(1.9, 19.7) | 6.9<br>(5.7, 8.9) | 0.9 |
| Died before discharge | 1 (3) | 0 | 0.4 |

\*For Acute COVID-19, n=5 patients identified as Asian and n=1 did not specify their race. For MIS-C, n=1 did not specify their race.
\*\*Other underlying conditions include active or prior oncologic issues, non-oncologic immunosuppressive disorder, rheumatologic/autoimmune disorder, renal or urologic dysfunction.
#Respiratory support included non-invasive supplementary oxygen or mechanical ventilation.

**Figure 1.**
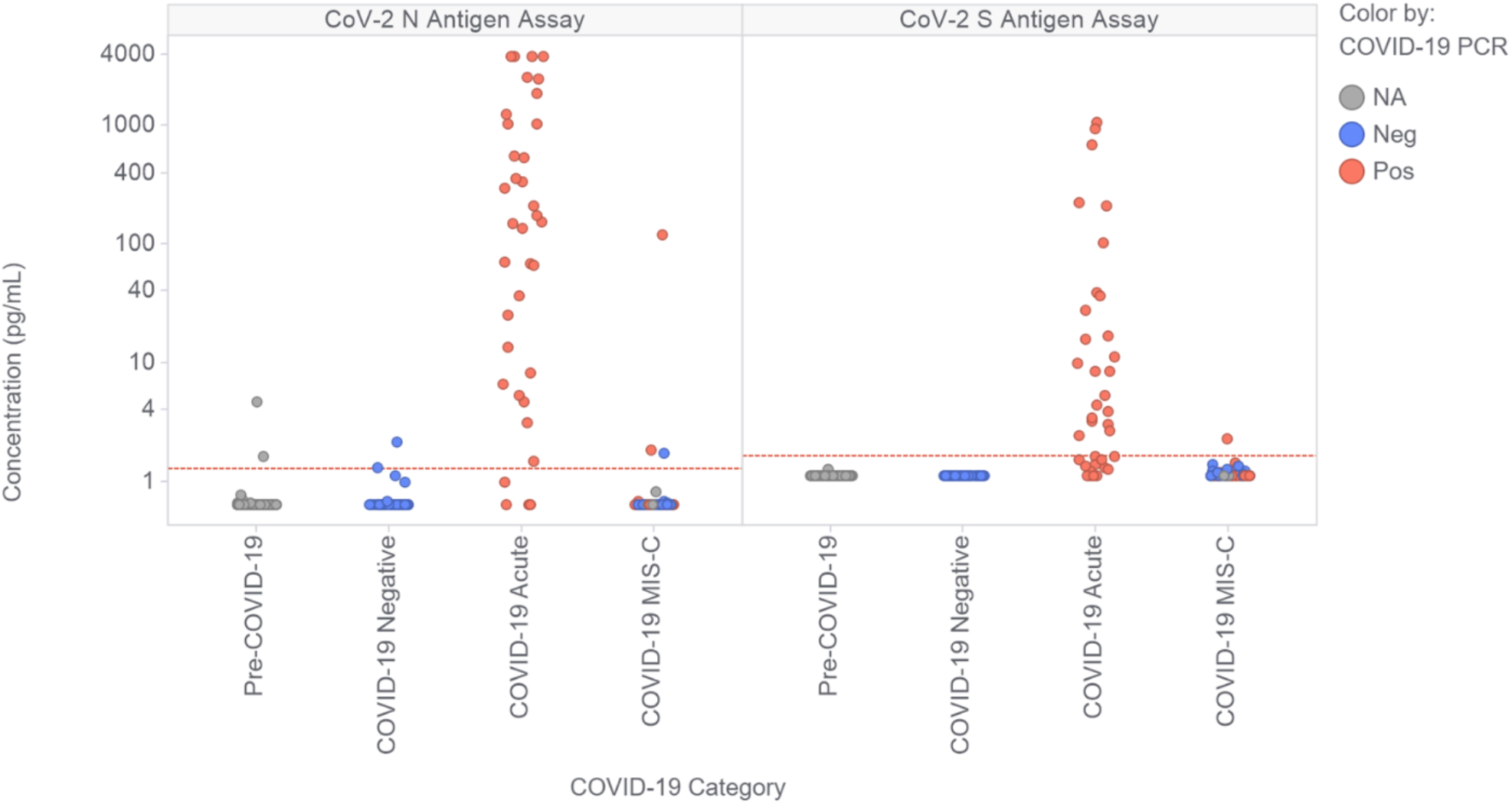
Measured levels of SARS-CoV-2 nucleocapsid (N) and spike (S) antigen in the plasma of children and young adult study participants. Participants were classified as controls with samples collected prior to 2020 (Pre-COVID-19, n = 67), controls ruled out for acute COVID-19 by negative nasopharyngeal swab RT-PCR (COVID-19 Negative, n = 43), patients with RT-PCR-confirmed acute COVID-19 infections (COVID-19 Acute, n = 36) and patients diagnosed as MIS-C (COVID-19 MIS-C, n = 53). Data points are colored based on the results of the most recent clinical COVID-19 RT-PCR test prior to sample collection. Of the MIS-C patients, the most recent clinical RT-PCR results prior to research blood sample collection were as follows: 20 RT-PCR+, 29 RT-PCR-, 4 NA (3 not performed, 1 inconclusive). The horizontal dashed red lines represent the assay thresholds for classifying samples as antigen positive.

In contrast, antigen concentrations in MIS-C patients [of whom 20 of 53 (38%) had a positive RT-PCR result at admission] were almost all undetectable. Concentrations of N ranged from < 1.28 pg/mL to 121 pg/mL, with only three (5.7%) samples above the cut-off value, two of which were from patients who were RT-PCR positive on admission. Concentrations of S ranged from < 1.65 pg/mL to 2.3 pg/mL, with only one sample above the cut-off value. The only MIS-C patient that had N concentration more than 5-fold above the assay cut-off (patient A, 121 pg/mL) was also the patient positive for S antigen, and had a positive clinical RT-PCR result 3 days prior. Of the 53 MIS-C patients, 35 had a research NP swab collected within 0-2 days of the blood sample (31/35 on day 0). Of those 35 swabs, 7 (20%) tested positive by RT-PCR. The patient with the lowest Ct value (22.8) was patient A; the other 6 RT-PCR positive patients all had Ct values >37. Of the 53 MIS-C patients, 40 had received IVIG prior to blood collection, including patient A. Of the 13 who had not received IVIG, none had detectable antigenemia.

The sensitivity and specificity of the N and S antigen assays in the study cohorts are shown in **Table 2**. The assays demonstrated high specificity in control patients. The specificity of the N assay was 97% in Pre-COVID-19 samples and 95% in COVID-19 Negative samples; S assay specificity was 100% in Pre-COVID-19 samples and 100% in COVID-19 Negative samples. The N assay sensitivity was 89% in all acute COVID-19 cases and 93% in cases with Ct value ≤ 35 on research NP swab RT-PCR. The S assay was less sensitive in acute COVID-19, with 64% sensitivity in all cases and 63% sensitivity in those with RT-PCR Ct values ≤ 35. Both assays were considerably less sensitive for identifying MIS-C, with sensitivity of 5.7% (N) and 1.9% (S), respectively.

**Table 2.**
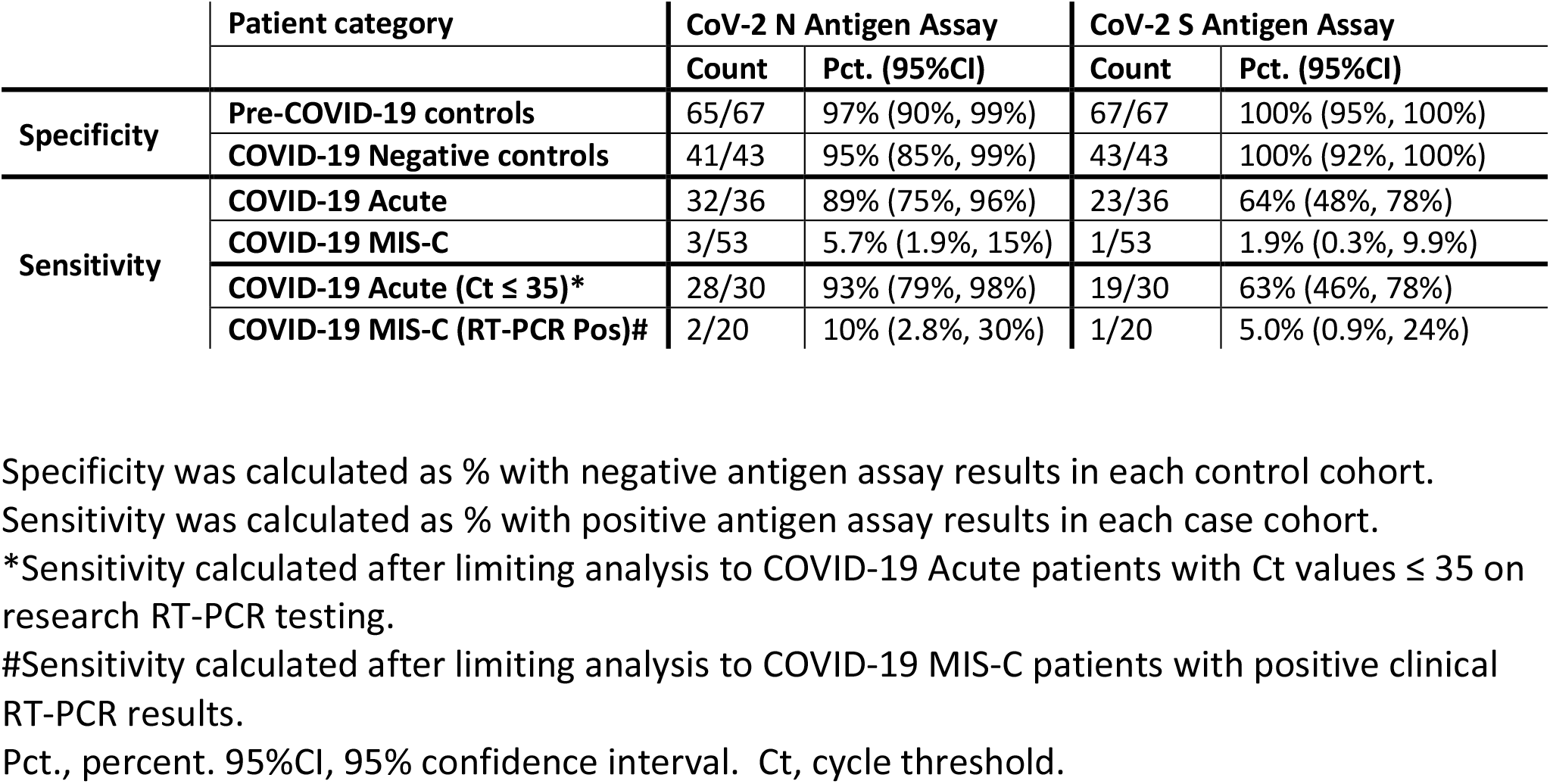
Sensitivity and specificity of assays for SARS-CoV-2 N and S antigens in plasma of pediatric patients with and without COVID-19.

|  | Patient category | CoV-2 N Antigen Assay |  | CoV-2 S Antigen Assay |  |
| --- | --- | --- | --- | --- | --- |
|  |  | Count | Pct. (95%CI) | Count | Pct. (95%CI) |
| **Specificity** | **Pre-COVID-19 controls** | 65/67 | 97% (90%, 99%) | 67/67 | 100% (95%, 100%) |
|  | **COVID-19 Negative controls** | 41/43 | 95% (85%, 99%) | 43/43 | 100% (92%, 100%) |
| **Sensitivity** | **COVID-19 Acute** | 32/36 | 89% (75%, 96%) | 23/36 | 64% (48%, 78%) |
|  | **COVID-19 MIS-C** | 3/53 | 5.7% (1.9%, 15%) | 1/53 | 1.9% (0.3%, 9.9%) |
|  | **COVID-19 Acute (Ct ≤ 35)\*** | 28/30 | 93% (79%, 98%) | 19/30 | 63% (46%, 78%) |
|  | **COVID-19 MIS-C (RT-PCR Pos)#** | 2/20 | 10% (2.8%, 30%) | 1/20 | 5.0% (0.9%, 24%) |

Specificity was calculated as % with negative antigen assay results in each control cohort.
Sensitivity was calculated as % with positive antigen assay results in each case cohort.
\*Sensitivity calculated after limiting analysis to COVID-19 Acute patients with Ct values ≤ 35 on research RT-PCR testing.

### Correlation of SARS-CoV-2 S and N Antigens with RT-PCR Ct values and Disease Severity

**Figure S1** shows the correlation of the measured S and N antigen concentrations in plasma with RT-PCR Ct values from the corresponding research NP swabs collected from acute COVID-19 patients. The correlations are relatively weak, with R^2^ values of 0.17 and 0.083 for N and S, respectively. The slopes are also much lower than would be expected for a linear dependence of plasma antigen concentration on respiratory RNA levels.

**Figure S2** examines the association of plasma N and S antigen concentrations with indicators of disease severity in acute COVID-19 patients. Admission to the ICU was associated with increased median (IQR) concentrations of both N [547.5 (53.0-2590.2) vs 51.6 (6.3-166.0) pg/mL, p=0.0009] and S [8.8 (2.3-130.6) vs 2.1 (1.4-4.0), p=0.002] antigens in plasma. Similarly, requiring any respiratory support (versus no respiratory support) was also associated with increased concentrations of N [320.0 (27.0-2130.3) vs 36.8 (4.5-160.2), p=0.003] and S [8.4 (1.90-86.0) vs 1.8 (<1.65-3.9), p=0.004], although the specific level of respiratory support (mechanical support vs. non-invasive supplemental oxygen) did not generate statistically significant differences for either antigen.

Measured concentrations of N and S within each sample are compared in **Figure S3**. Log-transformed concentration values (for samples that provided concentrations above the cut-off values for both assays) are moderately correlated with an R^2^ value of 0.52. The median ratio (22, IQR: 5.2-51) of N to S indicates consistently higher N than S concentrations but high variability in the relative amounts of the two antigens. There were no samples in which S but not N was detected.

### Correlation of SARS-CoV-2 Antigen and Antibody Measurements

In addition to antigen testing, all samples were also tested using a multiplexed serologic assay (**Methods, Supplementary Methods, Figure S4**). **Figure 2(a)** shows measured concentrations of anti-N and anti-S antibodies in the four patient cohorts from Figure 1. As expected, antibody levels in control samples were low: levels were below assay cut-offs for N and S antibodies in 100% (67/67) and 99% (66/67) of pre-COVID-19 samples and 99% (42/43) and 77% (33/43) of COVID-19 negative samples. All 10 COVID-19-negative control samples classified as positive by serology for S were also positive for anti-RBD activity (**Figure S4**), and likely represent prior vaccination or infection (data were not available for most COVID-19 negative patients, but the sample with highest anti-S level was from a vaccinated patient). Of the acute COVID-19 patients (all of whom were unvaccinated), 22% (8/36) and 36% (13/36) of samples were above the cut-off values for N and S antibodies. Nearly all MIS-C patients had seroconverted, with 94% (50/53) and 98% (52/53) above the N and S cut-off values.

**Figure 2.**
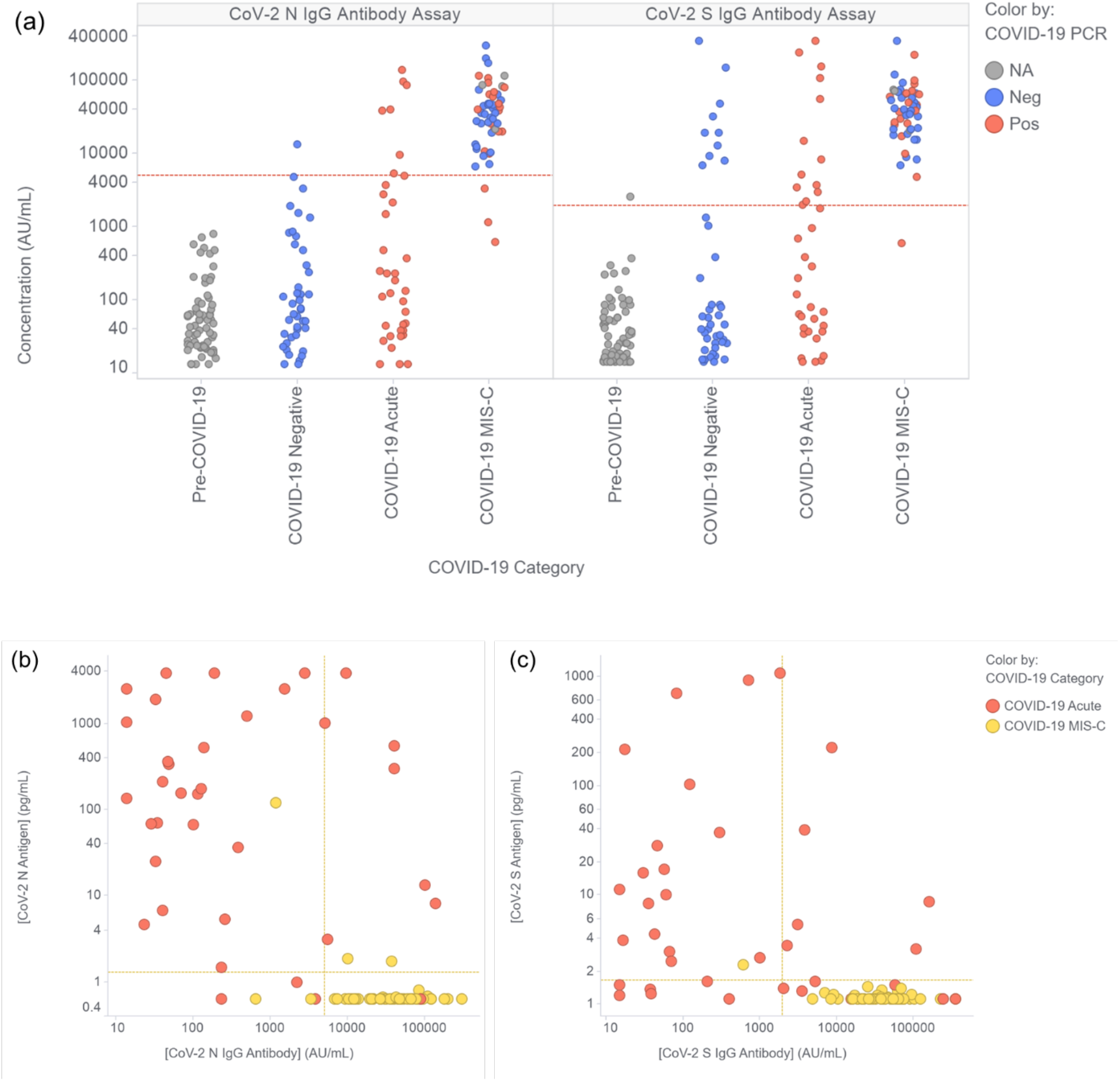
Measured levels of plasma IgG antibodies against SARS-CoV-2 N and S antigens and correlation with concentrations of N and S antigens. (a) Measurement of IgG antibodies against SARS-CoV-2 nucleocapsid (N) and spike (S) antigen in the plasma of children and young adult study participants. Participants were classified as described in Figure 1. Data points are colored based on the results of the most recent clinical COVID-19 RT-PCR test prior to sample collection. The horizontal dashed red lines represent the assay thresholds for classifying samples as antibody positive (Supplementary Methods). (b) and (c) Correlation of the levels of antigen and anti-antigen antibodies for SARS-CoV-2 N (b) and S (c) for the data points categorized as COVID-19 Acute or COVID-19 MIS-C in Figures 1 and 2a. Horizontal and vertical dashed yellow lines represent the assay thresholds for classifying samples as antigen or antibody positive, respectively.

**Figures 2(b)** and **2(c)** compare N and S antigen concentrations with antibody concentrations in each sample. For the acute COVID-19 samples, seroconversion had no apparent effect on the detection of N antigen, with N detected in 89% (25/28) of seronegative and 88% (7/8) of seropositive samples, and led to a small decrease in the detection of S antigen, with S detected in 74% (17/23) of seronegative samples, and 46% (6/13) of seropositive samples.

## Discussion

Our results using the MSD® S-PLEX CoV-2 N and S assays demonstrate that early in the hospital course, SARS-CoV-2 nucleocapsid and spike antigens are detectable in the blood of most pediatric patients with acute COVID-19, but in few patients with MIS-C. Specificity of both N and S assays was near 100% in samples from pre-COVID-19 and COVID-19-negative controls. Blood concentrations of N and S antigens in acute COVID-19 correlated with disease severity as indicated by ICU admission and need for respiratory support, but did not strongly correlate with RT-PCR Ct values in temporally matched NP samples. In contrast, antigen levels measured in NP swab samples have been shown to correlate closely with Ct values in NP samples [3, 4], suggesting that antigen levels in blood may be influenced by additional factors, such as infection in other tissues or variable antigen clearance from blood.

In acute COVID-19 patients, sensitivity of the S-PLEX blood N antigen assay was 89% overall and 93% if the corresponding NP sample Ct was ≤ 35, consistent with prior findings in adults with acute COVID-19. Shan *et al* found that a Simoa assay for N antigen in blood had 97.5% positive and 100% negative agreement with nasopharyngeal RT-PCR [7]. Wang *et al*, applying the S-PLEX N antigen assay to plasma of adults with acute COVID-19, demonstrated 91.9% clinical sensitivity (antigen-negative samples belonged to patients with high respiratory sample Ct values) and 94.2% clinical specificity [8]. Notably, they observed plasma N concentration ranges and association with disease severity similar to what we observed in children. The S-PLEX spike assay targets the receptor binding domain (RBD) within the S1 subunit; it can capture either the S1 domain (created by proteolytic cleavage at the S1-S2 junction [14]) or the full-length extracellular S1-S2 domain. Our spike assay had lower clinical sensitivity than our N assay in pediatric patients with acute COVID-19 (64% vs 89%), likely due to the consistently lower concentrations of S relative to N (approximately 22-fold).

The clinical overlap between MIS-C and toxic shock syndrome led to the hypothesis that SARS-CoV-2 spike peptides might function as a superantigen, contributing to T cell activation and MIS-C. [15, 16]. Yonker *et al* [9] tested the hypothesis that persistent SARS-CoV-2 infection in the gastrointestinal tract leads to antigenemia, potentially underlying MIS-C. Using Simoa-based assays previously developed and applied to the plasma of adults with acute COVID-19 by Ogata *et al* [6], they found higher signals from S1-S2, S1, and N assays in blood of MIS-C patients (medians of approximately 70, 50, and 5 pg/mL, respectively) relative to children with acute COVID-19 and pre-pandemic controls, and identified SARS-CoV-2 RNA in stool in 7/12 patients. They reported no difference in blood antigen concentrations between healthy pre-COVID-19 controls and acute COVID-19 patients.

We were unable to confirm the findings of the single-center study by Yonker *et al* [9]. One possible explanation may be differences in the clinical cohorts studied. Our study is larger and includes patients enrolled across the United States with MIS-C and with a range of acute COVID-19 disease severity, including those requiring mechanical ventilation and one that died (Table 1). Blood collection in our patients was performed at a single timepoint soon after admission; we did not perform serial sampling nor attempt to associate antigen levels with detection of virus in stool samples. The assays deployed in the two studies utilize different antibodies, so it is possible that S-PLEX assays were less efficient in detecting antigens in the blood of MIS-C patients. However, we reliably detected both N and S in children with acute COVID-19, and the MSD N antigen assay has demonstrated strong performance in pediatric and adult NP samples [3, 4], adult saliva [10], and adult blood [8]. Loss of antigens in complexes with host antibodies is possible, but is also unlikely to explain fully the disparate findings, as we still detected antigen after seroconversion in some acute COVID-19 patients. Differences in assay specificity may also explain differences in detection.

Our study has several strengths. We utilized rigorously validated and commercially available ultrasensitive and quantitative assays for N and S antigens, with the N antigen assay having already demonstrated high sensitivity and specificity in multiple sample types, including blood from adults [3, 4, 8, 10]. Samples were drawn from a diverse and nationally-representative multi-site CDC study with carefully adjudicated cohorts of children and young adults with acute COVID-19 (with a range of severity) and MIS-C. These samples had clearly documented timing for both RT-PCR testing and blood sample collection, allowing analysis of MIS-C blood samples drawn close to a validated clinical RT-PCR test result for comparison. The samples were collected with careful attention to handling, with minimal opportunity for antigen degradation.

Our study also had several limitations. First, all acute COVID-19 patient samples were from hospitalized patients, and thus our results may or may not overestimate the sensitivity of the assay in children with mild COVID-19. Second, 75% of MIS-C patients received IVIG prior to collection of the blood sample used in this analysis, raising the question of whether IVIG may have interfered with antigen detection. However, we did not detect N or S antigens in any of the 13 MIS-C patients who had pre-IVIG sampling, and the two acute COVID-19 patients who received IVIG prior to blood collection both had high blood antigen concentrations. Moreover, the presence of SARS-CoV-2-specific antibodies did not appear to inhibit antigen detection in acute COVID-19 patients. The Yonker *et al* study similarly included many post-IVIG samples, and concluded that IVIG initiation did not seem to have an impact on S antigen levels measured on serial samples in the few patients who had pre-IVIG measurements [9].

In conclusion, in this multicenter representative cohort of U.S. children with acute COVID-19 or MIS-C, we demonstrate that blood SARS-CoV-2 antigen measurement may be useful for diagnosing hospitalized children with acute COVID-19. Our findings do not support the hypothesis that ongoing SARS-CoV-2 spike antigenemia is a major contributor to MIS-C pathogenesis.

## Data Availability

All data produced in the present study are available upon reasonable request to the authors

## Notes

### Author Contributions

G.B.S., A.M., and N.R.P. conceived and designed the assay evaluation and sample analysis, and A.G.R., T.N., and N.B.H. designed the Overcoming COVID-19 study. All members of the study group acquired the data. G.B.S., N.M., P.B, J.J., N. P., and D.R. acquired the S-PLEX data. A.G.R., T.N., L.L.L., S.P.S., T.C.W., J.C.F., K.M.T., M.S.Z., J.E.S., N.B.H., M.C., A. B. M., M.A. S., K. I., H.R. F., B. M. C., H. C., and S. J. G. provided samples and clinical data from the Overcoming COVID-19 Immunobiology Study. G.A.M., J.J., and Y.Z. acquired control samples. G.B.S. and N.R.P. analyzed and interpreted the data and drafted the manuscript. Critical revisions to the manuscript were made by all members of the study group. N.R.P. and A.G.R. obtained the funds for the study. G.B.S., T.N., Y.Z, A.G.R., and N.R.P. verified all data. All authors had full access to all the data in the study and had final responsibility for the decision to submit for publication.

## Acknowledgements

The authors would like to thank Dr. Mark Kellogg, Rebecca Sprague, Caitlin Barrett, and Kaitlyn Daugherty for assistance with control samples.

This work represents the findings and conclusions of the authors and not the U.S. Centers for Disease Control and Prevention.

