## Supplementary Material for "Measurement of SARS-CoV-2 antigens in plasma of pediatric patients with acute COVID-19 or MIS-C using an ultrasensitive and quantitative immunoassay"

**Overcoming COVID-19 Immunobiology Study Group Investigators**

**(listed in PubMed, and ordered by U.S. State)**

The following study group members were all closely involved with the design, implementation, and oversight of the Overcoming COVID-19 immunobiology study.

**Alabama:** Children's of Alabama, Birmingham. Michele Kong, MD; Meghan Murdock, RN.
**Arizona:** University of Arizona, Tucson. Mary Glas Gaspers, MD; Katri V. Typpo, MD; Connor P. Kelley, MPH.
**Arkansas:** Arkansas Children's Hospital, Little Rock. Ronald C. Sanders, MD; Glenda Hefley MNSC, CCRP; Ashlyn Madding, RN, BSN; Masson Yates, RN, BSN.
**California:** UCSF Benioff Children's Hospital Oakland, Oakland. Natalie Z. Cvijanovich, MD; Geraldina Lionetti, MD; Juliana Murcia-Montoya, BS; Annie Higa, BS.
**California:** UCSF Benioff Children's Hospital, San Francisco. Denise Villarreal-Chico, BA.
**Colorado:** Children's Hospital Colorado, Aurora. Aline B. Maddux, MD, MSCS; Emily Port, BA, PMP; Rachel Mansour, BSN, RN, CPN.
**Florida**: Holtz Children's Hospital, Miami. Brandon M. Chatani, MD. Dr. Gwenn E McLaughlin, MD
**Florida.** Nicklaus Children's Hospital Research Institute, Miami. Paula S. Espinal, MD, MPH; Joseph M. Dunn.
**Georgia:** Emory University School of Medicine and Children's Healthcare of Atlanta, Atlanta. Satoshi Kamidani, MD.
**Illinois:** Ann & Robert H. Lurie Children's Hospital of Chicago, Chicago. Kelly N. Michelson, MD, MPH, Jovita Rodrigues, Hassan Khan, Jairo Chavez, Avani Shukla
**Indiana**: Riley Children's Health, Indiana University, School of Medicine, Indianapolis. Courtney Rowan, MD; Marla Lott; Kirsten Ramberg; Betsy Tudor.
**Massachusetts:** Boston Children's Hospital, Boston. Margaret M. Newhams, MPH; Suden Kucukak, MD; Sabrina R. Chen, BS; Benjamin J. Boutselis; Cameron Young; Hye Kyung Moon, MA; Jeni Melo, BS; Megan Elkins, MHS; Takuma Kobayashi, BS.
**Michigan**: CS Mott Children's Hospital, Ann Arbor. Chaandini Jayachandran, MSCCRP; Nadine L. N. Halligan
**Minnesota:** University of Minnesota Masonic Children's Hospital, Minneapolis, Janet R. Hume, MD, PhD; Ellen R. Bruno, MS; Lexie A. Goertzen, BA.
**Minnesota:** Mayo Clinic, Rochester. Emily R. Levy, MD; Supriya Behl, MSc; Noelle M. Drapeau, BA.
**Mississippi**: University of Mississippi Medical Center, Jackson. Charlotte V. Hobbs, MD; Urita Agana, BS; Gurbaksh Singh, BS.; Kalya Patterson, BS.
**Missouri:** Children's Mercy Kansas City, Shannon M. Hill, RN, BSN; Abigail Kietzman, ACRP-CP.
**Nebraska:** Children's Hospital & Medical Center, Omaha. Rachel Wellman.
**New Jersey:** St. Barnabas Medical Center, Livingston. Shira J. Gertz, MD.

**North Carolina:** University of North Carolina at Chapel Hill, Chapel Hill. Miriam Davis, RN; Angelo G. Navas; Paris C. Bennett; Will Lovell.
**Ohio:** Rebecca D. Considine Research Institute, Akron Children's Hospital, Akron. Ryan A. Nofziger, MD; Nicole A. Twinem, RN, ADN;
**Ohio:** Cincinnati Children's Hospital, Cincinnati. Chelsea C. Rohlfs, BS, MBA.
**Ohio**: University Hospitals Rainbow Babies and Children's Hospital, Cleveland. Steven L. Shein, MD; Rajashri Rasal, MPH.
**Pennsylvania:** Children's Hospital of Philadelphia, Philadelphia. Kathleen Chiotos, MD, MSCE; Ryan Burnett, BS; Rebecca L. Douglas, RN, BSN.
**South Carolina:** MUSC Children's Health, Charleston. Elizabeth H. Mack, MD, MS; Megan M. Bickford, MS; Lauren E. Wakefield, RN; Andrew M. Atz, MD; Laura Smallcomb, MD.
**Tennessee:** Monroe Carell Jr. Children's Hospital at Vanderbilt, Nashville. Natasha B. Halasa, MD, MPH; Meenakshi Golchha, MD; Rendie McHenry.
**Texas:** Texas Children's Hospital (Baylor), Houston. Grace Wu.
**Utah**: University of Utah, Primary Children's Hospital, Salt Lake City. Krow Ampofo, MD, MBChB; Evan Heller, CCRC.

**Supplementary Methods**

Overcoming COVID Immunobiology Study details
Study sites relied on a single IRB at Boston Children's Hospital, and informed consent was obtained from at least one parent or legal guardian. All pediatric patients were < 21 years old with confirmed SARS-CoV-2 positive reverse transcription polymerase chain reaction (RT-PCR) or antibody testing. Patients with immune compromising conditions that could impair antibody responses were excluded, as were patients with life support limitations or end stage lung disease. Hospitalized patients with COVID-19-related complications were hospitalized for acute COVID-19 or multisystem inflammatory syndrome in children (MIS-C) as defined below. Research blood and nasopharyngeal swabs were collected as soon as possible after admission. Data collected included demographics, past medical history including any chronic health conditions, RT-PCR and antibody testing results, and treatments and hospital course. Research blood samples (Lavender EDTA vacutainers) were centrifuged at 1300 x g for 10 min at room temperature, aliquoted, and frozen (-80$^{o}$C) within 1 hour (maximum of 6 hrs).

- **Acute COVID-19** was defined as having signs or symptoms that could be associated with early SARS-CoV-2 infection accompanied by a positive RT-PCR test for SARS-CoV-2 as defined by the U.S. Centers for Disease Control and Prevention (CDC) as listed on their website ([17], approved April 5$^{th}$, 2020).
    - At least two of the following symptoms: fever (measured or subjective), chills, rigors, myalgia, headache, sore throat, new olfactory and taste disorder(s); AND
    - Respiratory symptoms: cough, shortness of breath, hypoxia or difficulty breathing; OR
    - Severe respiratory illness with at least one of the following:
        - Clinical or radiographic evidence of pneumonia, OR
        - Acute respiratory distress syndrome (ARDS).

      AND
    - No alternative more likely diagnosis causing the symptoms.
- **MIS-C patients** met the criteria for multisystem inflammatory syndrome in children as defined by the CDC [2] as listed on their website ([5], published 5/14/2021), including:
    - An individual aged <21 years presenting with fever*, laboratory evidence of inflammation**, and evidence of clinically severe illness requiring hospitalization, with multisystem (≥2) organ involvement (cardiac, renal, respiratory, hematologic, gastrointestinal, dermatologic or neurological); AND
    - No alternative plausible diagnoses; AND
    - Positive for current or recent SARS-CoV-2 infection by RT-PCR, serology, or antigen test; or exposure to a suspected or confirmed COVID-19 case within the 4 weeks prior to the onset of symptoms.

        *Fever ≥38.0°C for ≥24 hours, or report of subjective fever lasting ≥24 hours
        **Including, but not limited to, one or more of the following: an elevated C-reactive protein (CRP), erythrocyte sedimentation rate (ESR), fibrinogen, procalcitonin, d-dimer, ferritin, lactic acid dehydrogenase (LDH), or interleukin 6 (IL-6), elevated neutrophils, reduced lymphocytes, and low albumin.

MSD S-PLEX CoV-2 N and MSD S-PLEX CoV-2 S assay design

The MSD S-PLEX CoV-2 N and MSD S-PLEX CoV-2 S assay kits (Meso Scale Discovery, Rockville, MD) employ a sandwich immunoassay format and electrochemiluminescence (ECL) detection. The assays are carried out in specially designed 96-well plate consumables (MSD Small Spot SECTOR™ plates) having integrated screen-printed carbon ink electrodes on the bottom of each well that are used as solid-phase supports for binding reactions, and as the source of electrical energy for inducing ECL from ECL labels in binding complexes on their surfaces. The kits use MSD's ultra-sensitive S-PLEX ECL format, which provides additional signal enhancement and sensitivity relative to conventional ECL formats.

The assays were run according to protocols described in the kit package inserts [11, 12]. Plasma samples were diluted 4-fold in the assay diluent provided with the kits prior to analysis. Sample quantitation was achieved by calibration of the assay using a calibration curve generated using a recombinant antigen standard, and fit to a four parameter logistic (4PL) model. For graphing and analysis, any concentrations below the limit of detection (LOD) were assigned the LOD value, and any concentrations above the highest calibration standard were assigned the concentration of the highest calibration standard.

The N antigen assay [11] utilizes recombinant full-length N-protein as a standard, and monoclonal capture/detection antibodies generated by MSD against the full-length recombinant N protein. The limit of detection (LOD) and assay cut-off for the N assay were previously established as part of a study of respiratory samples tested without additional dilution [3]. For the present study, the previously established LOD (0.16 pg/mL) and cut-off values (0.32 pg/mL) were multiplied by four (LOD = 0.64 pg/mL, cut-off = 1.28 pg/mL) to account for the 4-fold sample dilution used for plasma samples.

The S antigen assay [12] utilizes recombinant receptor binding domain (RBD) of the S1 subunit of the spike protein as a standard, and monoclonal capture/detection antibodies generated by MSD against recombinant RBD. The LOD for the S assay was determined as the concentration (based on a 4PL fit to a calibration curve) that provides a signal 2.5 standard deviations above the blank signal. The measured LOD value was 0.28 pg/mL or 1.12 pg/mL after adjustment for the 4-fold dilution of plasma samples. The cut-off for the S assay was determined as the concentration that was 2.5 standard deviations above the average concentration measured for a set of 24 pre-COVID adult plasma samples. The determined cut-off for the S antigen assay was 0.412 pg/mL in the diluted samples, or 1.65 pg/mL after adjustment for the 4-fold plasma dilution.

SARS-CoV-2 Antibody (Serology) Assays

Multiplexed measurements of IgG antibodies against a set of antigens from SARS-CoV-2 and other coronaviruses were conducted in an indirect serology format with ECL detection using the MSD V-PLEX® COVID-19 Coronavirus Panel 2 Kit (Meso Scale Discovery, Rockville, MD) [13]. The assays are carried out in 96-well plate consumables (MSD 10-Spot SECTOR plates) having integrated screen-printed carbon ink electrodes, as described for the antigen assays, except that the electrodes are configured to support immobilized arrays of up to 10 antigens within each well. In the indirect serology format, samples are incubated in the wells to allow antibodies to bind to the immobilized array of antigens. After washing out the sample, a monoclonal anti-human IgG antibody is incubated in the well to bind to any capture IgG and allow for ECL detection. The assays were run according to the protocol provided with the assay kit. Sample were diluted 5000-fold in the assay diluent provided with the kits prior to analysis.

The kit was used to measure antibody responses to an array of nine recombinant coronavirus antigens. These included four SARS-CoV-2 antigens: Nucleocapsid (N), Spike (S), the Receptor Binding Domain (RBD) fragment of the spike protein and the N-Terminal Domain (NTD) of the spike protein. The array also included the S protein from five additional coronaviruses: SARS-CoV-1, 229E, HKU1, NL63 and OC43. All of the S protein constructs were expressed as fusions of the spike ectodomain with the T4 phage foldon domain, so that the spike ectodomain assumes its native trimeric structure. Sample quantitation was achieved by calibration of the assay using a calibration curve generated using a pooled convalescent COVID serum standard, and fit to a four parameter logistic (4PL) model. Quantitation is expressed in arbitrary units per mL (AU/mL). Concentration values for SARS-CoV-2 N, Spike, and RBD have been referenced to the WHO COVID-19 serology standard [13]. The conversion factors for converting from MSD AU/mL to WHO Binding Antibody Units per mL (WHO BAU/mL) are 0.00236 (N), 0.00901 (Spike) or 0.0272 (RBD). Cut-off concentration values for identifying seroconversion based on the SARS-CoV-2 N (5,000 AU/mL), S (1,960 AU/mL) and RBD (538 AU/mL) antigens were determined from a study of 200 adult pre-COVID serum samples and 176 adult convalescent COVID serum samples [13].

<u>Data and Statistical Analysis</u>

For analysis of clinical data, Pearson Chi-Square or Fisher's exact test (2-sided) (applied if <5 counts) were used for categorical data, and Mann Whitney U with Tukey's median IQR was used for numerical data.

For all analysis related to antigen concentrations, data and statistical analysis were carried out using the R statistical programing language (Version 4.0.3). Linear regression used the "lm" function in R. Comparison of groups was conducted with the non-parametric one-sided Mann-Whitney test using the "wilcox.test" function in R. Confidence limits for proportions were calculated using the "prop.test" function in R. Graphs were generated using TIBCO Spotfire software (version 11.2.0.59).

**Supplementary Results**

Table S1 summarizes key laboratory and blood sample handling data for COVID-19 Acute and MIS-C patients.

Figure S4 shows the measured concentrations of antibodies against all the antigens included in the serology test panel. Antibody activity against the SARS-CoV-2 spike subunit antigens (RBD and NTD), and the SARS-CoV-1 spike (which has high homology to the SARS-CoV-2 spike) show similar behavior as described in the main results section for activity against the full SARS-CoV-2 spike antigen (see discussion of Figure 2). Interestingly, the activity against spike from the four common pre-COVID circulating coronaviruses was significantly higher ($p < 0.0001$) for the COVID-19 MIS-C group than each of the other study categories. Relative to the Pre-COVID-19 samples, the COVID-19 MIS-C samples had median levels for the circulating coronavirus spike antigen that ranged from 4-fold higher (NL63) to 10-fold higher (229E). The increases in median levels for COVID-19 MIS-C relative to COVID-19 Acute ranged from 3-fold higher (NL63) to 6-fold higher (229E).

**Table S1. Laboratory and Blood Specimen Handling Data**

| **Specimen Handling** | **COVID-19 Acute n=36** | **COVID-19 MIS-C n=53** | **P-value** |
|---|---|---|---|
| **SARS-CoV-2 RT-PCR+** (clinical test, performed prior to blood draw) n (%) | 36 (100) | 20 (38) | **<0.0001** |
| **Ct value for N1/N2 target**[#] (research NP swab) Median (IQR) | 27.2 (24.4, 34.1) | 37.7 (37.2, 38.5) | **0.004** |
| **SARS-CoV-2 Antibody+** (clinical test) n (%) | 5 (14) | 48 (91) | **<0.0001** |
| **Hours between research blood collection, processing, and freezing sample** Median (IQR) | 1.1 (0.8, 1.5) | 1.1 (0.8, 1.4) | 0.8 |
| **Days between research blood and research NP swab collection*** Median (Range) | 0 (0 – 1) | 0 (0 – 2) | 0.6 |
| **Days between hospital clinical RT-PCR test and research blood** Median (IQR) | 2 (1, 3) | 2** (1, 4) | 0.3 |

Ct, cycle threshold. IQR, interquartile range.
### Ct values presented are average of N1 and N2 targets. All acute COVID-19 patients had research NP swabs collected within 1 day of blood sample. Of the 53 MIS-C patients, 35 had a research NP swab collected within 0-2 days of the blood sample (31/35 on day 0). Of those 35 swabs, 7 tested positive by RT-PCR and generated a Ct value.
*relevant for analysis of blood antigen vs NP Ct value
** RT-PCR test not conducted for 3 MIS-C patients
Pearson Chi-Square or Fisher's exact test (2-sided) applied if <5 counts used for categorical data.
Mann Whitney U with Tukey's median IQR was used for numerical data.

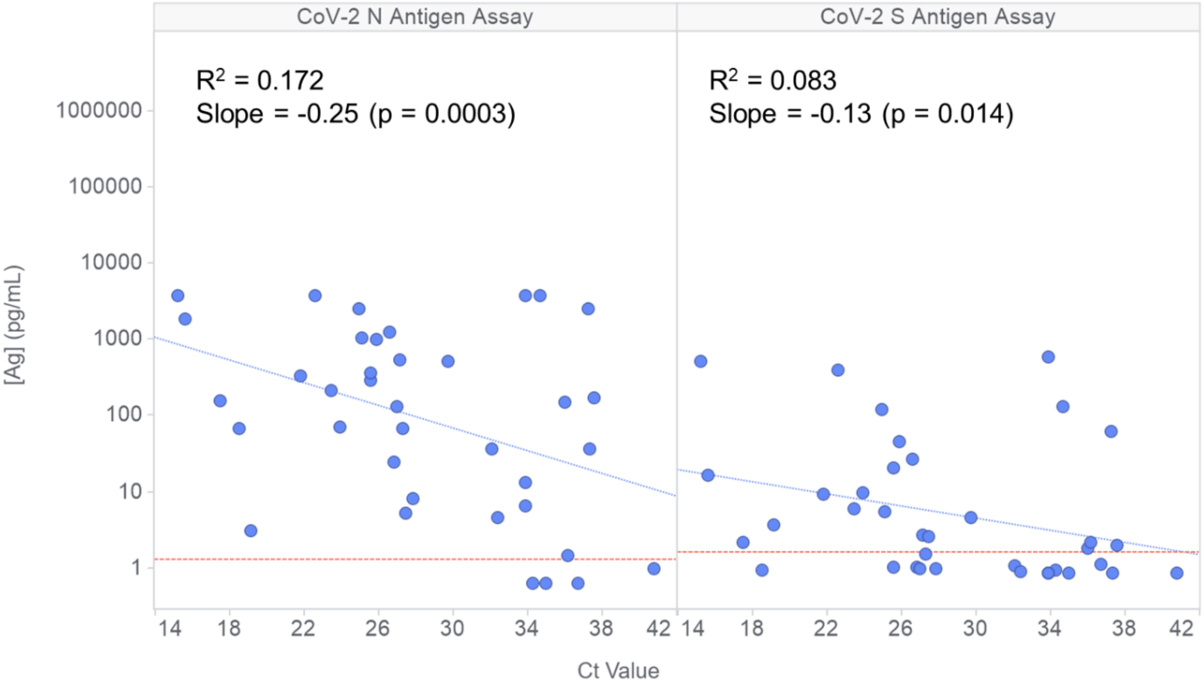


Figure S1. Correlation of SARS-CoV-2 nucleocapsid (N) and spike (S) antigen concentrations in plasma with SARS-CoV-2 RT-PCR cycle threshold (Ct) values measured in NP swab samples collected within 24 hours of the plasma sample in patients with severe acute COVID-19. The y and x axis show the log of the measured plasma antigen concentrations vs. the Ct value from swab RT-PCR testing. Linear regression using the log2-transformed values for the antigen concentrations provided the linear fits indicated by the blue dashed lines, and the $R^2$ and slope values provided within each panel. A perfect correlation would be expected to give a slope of around -1.0. The red horizontal dashed lines represent the assay threshold values for classifying samples as antigen positive.

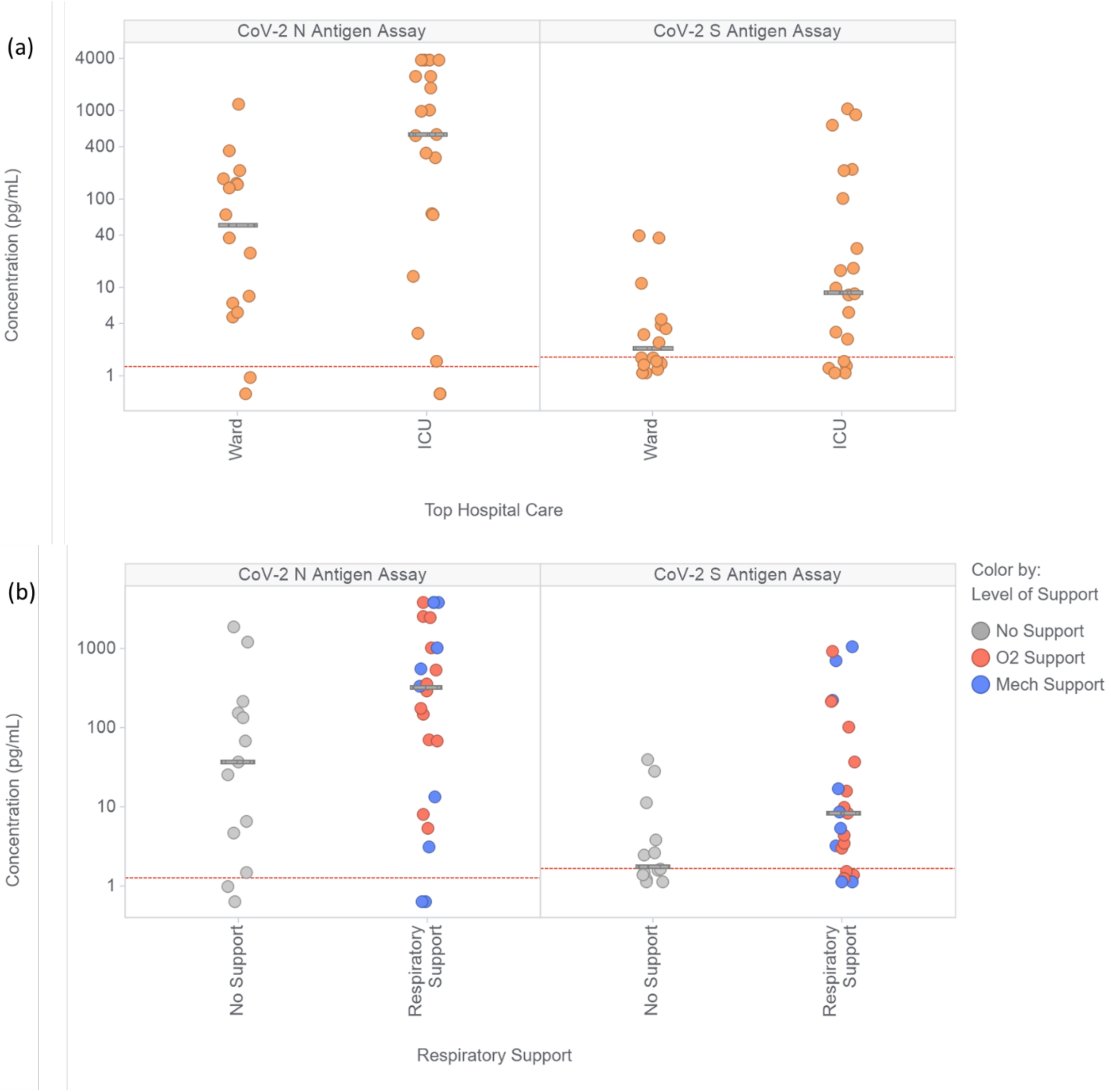


<u>Figure S2.</u> Association of measured plasma concentrations in samples from acute COVID-19 patients with (a) level of hospital care and (b) level of respiratory care (no support, grey dots; supplementary oxygen provided non-invasively, red dots; or invasive mechanical support (endotracheal tube or tracheostomy), blue dots). The red dashed lines indicate the assay thresholds for each assay for classifying samples as antigen positive. The median value for each category is indicated with a grey bar.

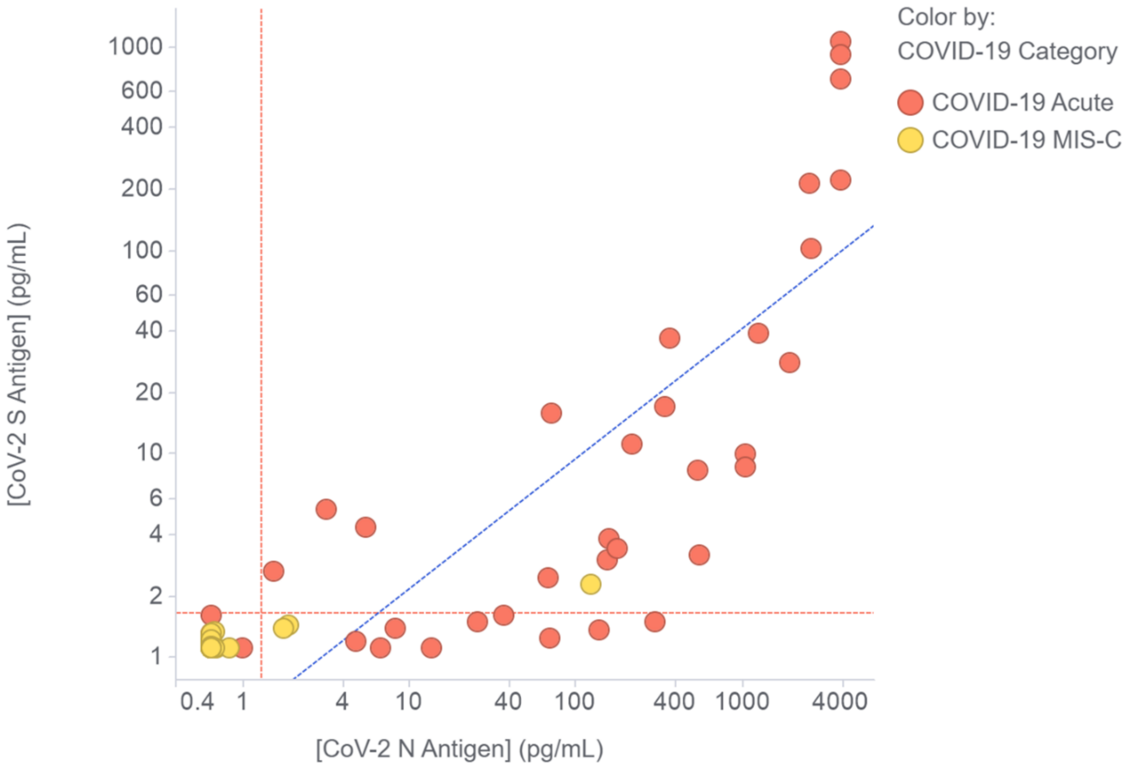


Figure S3. Correlation of SARS-CoV-2 spike (S) antigen with nucleocapsid (N) antigen in the plasma of children and young adults with acute COVID-19 (red) or MIS-C (yellow). The red dashed lines indicate the assay thresholds for each assay for classifying samples as antigen positive. The blue dashed line shows the linear regression fit to the data points that are above the assay thresholds for both assays.

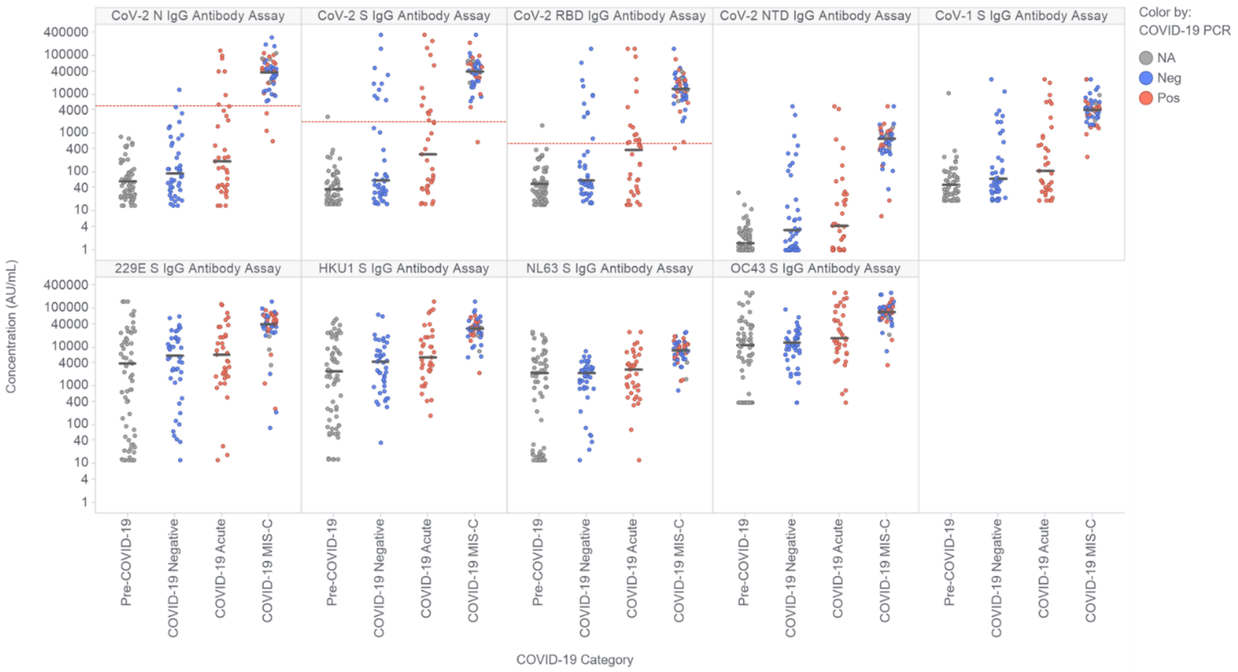


Figure S4. Measurement of levels of IgG antibodies against nucleocapsid (N), spike (S), spike receptor binding domain (RBD) and spike N-terminal domain (NTD) antigens of SARS-CoV-2, and the spike antigen of SARS-CoV-1 and four pre-COVID-19 common circulating coronaviruses (229E, HKU1, NL63, OC43). Participants were classified as described in Figure 1. Data points are colored based on the results of the most recent clinical COVID-19 RT-PCR test prior to sample collection. The black bars indicate the median value for each group. The horizontal dashed red lines represent the assay thresholds for classifying samples as antibody positive for the SARS-CoV-2 nucleocapsid, spike and RBD antigens. Thresholds were not established for the other antigens.
